# Pharmacokinetic /Pharmacodynamic Considerations of Alternate Dosing Strategies of Tocilizumab in COVID-19

**DOI:** 10.1101/2021.08.28.21262692

**Authors:** Elizabeth Leung, Sarah C.J. Jorgensen, Ryan L. Crass, Sumit Raybardhan, Bradley Langford, W. Justin Moore, Nathaniel J. Rhodes

**Affiliations:** Department of Pharmacy, St. Michael’s Hospital/Unity Health Toronto, Toronto, ON, Canada; Leslie Dan Faculty of Pharmacy, University of Toronto, Toronto, ON, Canada; Li Ka Shing Knowledge Institute, Toronto, ON, Canada; Department of Pharmacy, Mount Sinai Hospital, Toronto, ON, Canada; Ann Arbor Pharmacometrics Group, Ann Arbor, MI, USA; Department of Pharmacy, North York General, Toronto, ON, Canada; Public Health Ontario, Toronto, ON, Canada; Department of Pharmacy, Northwestern Medicine, Chicago, IL, USA; Department of Pharmacy Practice, College of Pharmacy Downers Grove Campus, Midwestern University, IL, USA; Pharmacometrics Center of Excellence, College of Pharmacy Downers Grove Campus, Midwestern University, IL, USA

**Keywords:** coronavirus, interleukins, pharmacokinetics, pharmacodynamics, pharmacology, monoclonal antibodies, immunopharmacology, SARS CoV-2, tocilizumab

## Abstract

Tocilizumab is one of few treatments that have been shown to improve mortality in patients with COVID-19, but increased demand has led to relative global shortages. Recently, it has been suggested that lower doses, or fixed doses, of tocilizumab could be a potential solution to conserve the limited global supply while conferring equivalent therapeutic benefit to the dosing regimens studied in major trials. The relationship between tocilizumab dose, exposure, and response in COVID-19 has not been adequately characterized. There are a number of pharmacokinetic (PK) parameters which likely differ between patients with severe COVID-19 and patients in whom tocilizumab was studied during the FDA approval process. Likewise, it is unclear whether a threshold exposure is necessary for tocilizumab efficacy. The safety and efficacy of fixed versus weight-based dosing of tocilizumab has been evaluated outside of COVID-19, but it is uncertain if these observations are generalizable to severe or critical COVID-19. In the current review, we consider the potential advantages and limitations of alternative tocilizumab dosing strategies. Leveraging PK models and simulation analyses, we demonstrate that a fixed single dose of tocilizumab 400 mg is unlikely to produce PK exposures equivalent to those achieved in the REMAP-CAP trial, though weight-stratified dosing appears to produce more uniform exposure distribution. Data from current and future trials could provide PK/PD insight to better inform dosing strategies at the bedside. Ultimately, rational dosing strategies that balance available limited supply with patient needs are required.

## Introduction/Background

Tocilizumab, a monoclonal antibody that inhibits interleukin-6 (IL-6) signaling by binding to soluble and membrane-bound IL-6 receptors (IL-6R), has been shown to improve mortality in patients with severe or critical COVID-19 [1,2]. It is currently recommended by a number of international, national, and local consensus guidelines for treatment of severely and critically ill COVID-19 patients, driven largely by the results of two major platform randomized controlled trials in COVID-19 [3,4]. Tocilizumab dosing strategies in these trials included banded, weight-based dosing (e.g., 400 mg if 41-65 kg, 600 mg if 66-90 kg, 800 mg if >90 kg), and weight-based dosing (e.g., 8mg/kg up to 800 mg)[3,4,5,6,7]. However, in the absence of exposure-response data to guide dose selection for patients with COVID-19, the optimal tocilizumab dosing strategy in this population remains unclear.

Recently, increased demand coupled with limited production capacity has led to a relative global tocilizumab shortage [8]. Various jurisdictions have had to balance high demand with limited supply [9,10,11,12]. Unfortunately, this has disproportionately impacted areas of the world already struggling with a large burden of cases. In light of the current tocilizumab shortage, some jurisdictions have implemented lower, fixed doses of tocilizumab as a rationing strategy with the hope that this will not compromise the observed benefit [8,9,10,11,13].

In this article, we review the pharmacokinetics (PK) and pharmacodynamics (PD) of tocilizumab and enumerate potential advantages and limitations of a fixed-dose approach for the treatment of COVID-19. We consider direct clinical experience as well as extrapolated evidence in other patient populations and conditions. We also consider whether fixed-dosing is likely to produce less interpatient variability in exposures across the body-weight distribution of adult patients compared to weight-based dosing. Finally, we present the results of model-based simulations to evaluate whether alternative dosing strategies for tocilizumab are likely to yield PK exposures that are substantially similar to those achieved in clinical trials conducted in COVID-19 patients.

### Overview of tocilizumab PK

The relationship between tocilizumab dose, exposure, and response in COVID-19 has not been adequately characterized. Weight-based dosing of tocilizumab in COVID-19 was adopted empirically based on experience in treating patients with rheumatological diseases and cytokine release syndrome (CRS) associated with chimeric antigen receptor T-cell (CAR T-cell) therapy. However, the PK properties of monoclonal antibodies leave open the possibility that fixed-dosing may reduce interpatient variability in exposure. Such an approach could hypothetically facilitate the use of fixed doses without compromising efficacy or safety.

Due to their large size and hydrophilicity, monoclonal antibodies reside almost exclusively within plasma and extracellular fluid [14], and these fluid compartments frequently do not scale linearly with incremental increases in total body weight [15,16,17,18]. Monoclonal antibodies are eliminated via two pathways. First, after administration, they undergo proteolytic catabolism, which is associated with linear clearance (CL). The second pathway involves target-engagement (i.e., IL-6R and tocilizumab) followed by internalization and intracellular degradation. This process is associated with non-linear clearance [14] and depends on the relative expression of the target (e.g., IL-6R, sIL-6R). Therefore, this mechanism can be influenced by patient-specific factors such as disease severity [14,19,20]. Notably, the linear portion of tocilizumab clearance appears to scale with body weight [14], but this relationship is imperfect [18,21].

As a result, weight-based dosing of monoclonal antibodies, which linearly proportionalizes the dose to total body weight (as opposed to weight-banded or allometric dose scaling), may result in relative under-exposure in low body weight patients and relative over-exposure in obese patients compared to patients with mid-range body weights [18,22]. In order to balance the need for appropriate dosing in severe COVID-19, the limited supply of tocilizumab available in some areas, and the potential risk of over- or under-exposure within different body-weight strata, a formal comparison of weight-based and alternative dosing strategies is needed.

### Evaluation of tocilizumab population PK in approved indications

Tocilizumab was initially approved by the Food and Drug Administration (FDA) Center for Drug Evaluation and Research (CDER) for the treatment of rheumatoid arthritis (RA) in 2010 and has since been approved in over 75 countries for RA and related conditions. The approved intravenous (IV) dose is 4 mg/kg every 4 weeks which can be increased to 8 mg/kg based on clinical response [23]. Frey et al. performed a population PK analysis using pooled data from four Phase 3 clinical trials in RA, and identified a two-compartment structural model with parallel linear and non-linear (Michaelis-Menten) elimination [24]. The non-linear portion of tocilizumab clearance following administration is concentration-dependent. Accordingly, the half-life varies based on the dose administered. The non-linear portion of clearance is believed to reflect target-mediated drug disposition after binding to the IL-6R [14,24]. In the initial population PK analysis submitted to the FDA, body surface area (BSA) was identified as a significant predictor of total clearance [21]. However, the relationship between tocilizumab exposure and body size was non-linear; therefore, predicted tocilizumab exposures were not matched across the body weight distribution with weight-based dosing (e.g., 8 mg/kg) [21]. Tocilizumab exposure (e.g., AUC and C_max_) was predicted to increase with increasing body weight with an 8 mg/kg dose; however, a fixed 560 mg dose led to slightly lower exposures at high body weight compared to more normal body weights. Ultimately, the weight-based dosing studied in tocilizumab clinical development was retained despite inadequate exposure matching across the weight range, due to the absence of a clear safety signal and a lower clinical response rate for high body weight RA patients, despite higher relative exposures [21].

Subsequently, a fixed-dose subcutaneous dosing regimen was studied and approved for RA in 2013. The approved subcutaneous (SC) dose was 162 mg every one to two weeks with weekly dosing recommended initially in patients weighing ≥100 kg [25]. A second population PK model was developed using data from two Phase 3 trials wherein both IV and SC dosing were evaluated [25]. This model was identical in terms of compartmental structural and parallel linear and non-linear elimination with the exception that an absorption parameter (k_a_) was added to accommodate for the SC route [25]. In contrast to the first model, this population PK model scaled all clearance (e.g., systemic and inter-compartmental) and volume (e.g., central and peripheral) parameters to total body weight. Given the marginal decrease in exposures (higher clearance) observed in patients with higher body weights, a weekly subcutaneous fixed dose (rather than every two weeks) was selected for this body-weight subgroup. Saturation of non-linear clearance was observed with an 8 mg/kg IV dose given every four weeks and with a fixed dose of 162 mg SC given every week, suggesting that fixed dosing is possible, so long as concentrations remain above the saturation threshold for non-linear clearance [25].

Tocilizumab was subsequently approved for polyarticular juvenile idiopathic arthritis (PJIA) and systemic juvenile idiopathic arthritis (SJIA). The approved doses of IV tocilizumab are 10-12 mg/kg (< 30 kg) and 8 mg/kg (≥30 kg) for these indications, respectively [23]. The population PK model supporting the dose selection for these pediatric patients was structurally identical to the initial RA model; however, height and age were included as additional covariates on the central distribution volume [26]. Similar to the initial IV tocilizumab population PK model, BSA was identified as a covariate on clearance with height and age identified as covariates of central volume [26]. The 8 mg/kg regimen was evaluated in the registrational study for pediatric patients with SJIA; however, it was noted that low body weight (<30 kg) pediatric patients had lower clinical response [26,27]. Modeling was used to predict individual exposures with higher dosing (12 mg/kg) for the low body weight patients and demonstrated good concordance with exposures achieved in the higher body weight range. However, based on these limited data and a lack of disease similarity, it is unclear whether these doses would extrapolate to pediatric patients with COVID-19.

The most recent tocilizumab indication, which may exhibit the greatest similarity to COVID-19, is for the treatment of severe or life-threatening CRS secondary to CAR T therapy [23,27]. FDA-approval for this indication was based on retrospective analysis of 45 adult and pediatric patients receiving tocilizumab for CRS in CAR T clinical trials (27). Patients received either 12 mg/kg (<30 kg) or 8 mg/kg (≥30 kg) as a single IV dose, which could be repeated. The key clinical pharmacology question in the FDA review of this CRS indication was the adequacy of the dosing regimen [27,28]. Exposure-response analysis was not possible as only a single dose-level was evaluated and only sparse PK data were available. Accordingly, the FDA utilized the previously developed pediatric model to support dosing in SJIA and re-estimated the linear portion of systemic clearance and central volume of distribution. All other PK parameters and inter-individual variation estimates were fixed at the previously determined values [27]. The typical values of linear systemic clearance and central distribution volume were found to be approximately three-fold and two-fold higher, respectively, in critically ill patients with severe or life-threatening CRS compared to patients with stable SJIA. In the absence of defined exposure-response for either efficacy or safety, the target exposure was defined based on the highest observed C_max_ (649 mg/L) with a single 28 mg/kg dose in five healthy subjects.

### Tocilizumab PK in patients with COVID-19

Tocilizumab PK data in patients with COVID-19 are currently limited to a single population PK analysis available as a pre-print [29]. Moes et al. conducted an open-label, single-center observational PK study of tocilizumab in 29 patients with severe COVID-19 who all received a dose of 8 mg/kg to a maximum of 800 mg. They collected 139 leftover laboratory samples for secondary use over a 20-day window, which were assayed to determine free tocilizumab and sIL-6R concentrations. C-reactive protein and clinical outcomes were also assessed. The final population model consisted of a single disposition compartment model with parallel first order and non-linear clearance. No significant covariate effects were identified. The estimates for CL and volume of the central compartment (Vc) were 0.725 L/day and 4.34 L. The linear CL estimate was higher than the estimate in adult RA patients (0.2-0.3 L/day), pediatric SJIA patients (0.17), and pediatric and adult patients with CAR T-induced CRS (0.5 L/day) although the central volume was largely consistent with other patient populations.

Simulations were performed to evaluate weight-based and fixed-dosing regimens with the conclusion that a 600 mg fixed dose may improve exposure matching across body weight; however, the limited sample precluded robust assessment of covariate effects (e.g., body size), which limits the external validity of conclusions about exposure variation with body weight. It is noteworthy that clearance was high in this cohort and most consistent with pediatric and adult patients with CRS; however, inter-individual variation in clearance and volume was modest and more consistent with patients with stable inflammatory diseases.

### How do IL-6 and IL-6R expression compare between COVID-19 and other inflammatory conditions?

When extrapolating dosing from one disease state to another, a critical consideration is whether or not disease similarity exists between the conditions in question. IL-6 belongs to the glycoprotein 130 (gp130)-related family of cytokines, responsible for host inflammatory regulation as well as cell growth and differentiation [30]. The IL-6R is expressed as a membrane-bound receptor (containing the gp130 receptor subunit) on hepatocytes, certain leukocytes, and megakaryocytes [31]. Notably, IL-6 is distinct from other pro-inflammatory cytokines due to the natural soluble form of the IL-6R (i.e., sIL-6R) which is detectable in human serum during periods of inflammation or infection. The majority of soluble cytokine receptors are associated with antagonistic functions, while the sIL-6R serves an agonist function and facilitates cytokine signaling [31,32]. IL-6 signaling through the membrane-bound IL-6R receptor complex is referred to as classical IL-6R signaling, whereas IL-6 signaling through sIL-6R is referred to as IL-6 trans-signaling [31,32,33,34]. Trans-signaling may cause cells without a membrane bound IL-6R, including epithelial cells, smooth muscle cells, and endothelial cells, to have a pro-inflammatory response to IL-6 in critical illness [34]. In vitro studies have shown that IL-6 signal transduction can also be initiated by the IL-6/sIL-6R complex; therefore, both membrane-bound IL-6R and sIL-6R are capable of participating in IL-6-mediated cell signaling [35].

Serum concentrations of IL-6 are elevated in chronic immune-mediated inflammatory diseases, and IL-6 levels have been shown to correlate with disease activity in conditions such as RA even though the degree of IL-6 expression may vary by disease condition [35]. IL-6 concentrations in patients with COVID-19 patients are heterogeneous [36,37,38] but generally similar to concentrations observed in patients with RA. Patients with COVID-19 who experience higher IL-6 concentrations have more severe disease, and an increased risk of mortality [36,38,39,40]. This trend mirrors observations from critically ill patients with severe sepsis, where non-survivors had higher mean serum IL-6 levels compared to survivors [41]. In contrast, IL-6 concentrations are significantly lower in patients with COVID-19 compared to patients with CRS-associated with CAR T-cell therapy [13,37,42,43]. Likewise, IL-6 concentrations in patients with acute respiratory distress syndrome (ARDS) and septic shock are highly variable and depend on the sampling method and observation time point within the course of illness. For example, IL-6 concentrations may peak early in the setting of acute inflammation, and then decrease rapidly to undetectable levels [34,43,44]. Critically ill patients with COVID-19 generally present with elevated inflammatory markers (e.g. IL-6, sIL-6R, CRP) compared to those with mild to moderate illness [38,42,43,44]. Single cell RNA-sequencing (RNA-seq) analysis of peripheral blood failed to identify circulating cells as the source of IL-6 in COVID-19 [45,46]. This finding supports the hypothesis that cells within the alveolar space rather than the periphery may be the source of enhanced IL-6 expression in COVID-19. A prospective observational trial compared IL-6 expression within the lung of patients with COVID-19 pneumonia to patients with pneumonia not caused by COVID-19 [47]. Bulk RNA-seq analysis of flow cytometry-sorted alveolar macrophages, collected using bronchoalveolar lavage, revealed that IL-6 expression was not different between patients with COVID-19 pneumonia compared to patients with pneumonia due to other pathogens [47].

Although IL-6 concentrations have been historically used as a surrogate for clinical response in chronic inflammatory conditions, recent evidence suggests that sIL-6R may be a more informative marker of immune dysregulation and a better predictor of tocilizumab response [40, 48]. Correspondingly, sIL-6R may also be a better surrogate for clinical response with tocilizumab in COVID-19. The proposed rationale for evaluating sIL-6R response is that trans-signaling via sIL-6R may provide a more complete picture of clinically relevant IL-6 activity in vivo [48]. Nishimoto conducted a tocilizumab PK/PD study in patients with RA and Castleman Disease and found that “as long as free tocilizumab was detectable, sIL-6R was saturated and IL-6 signaling was completely inhibited” [35]. The threshold concentration for saturation was 1 μg/mL. However, even if sIL-6R is more predictive of tocilizumab anti-inflammatory response it is much more challenging to measure clinically due limited assay availability and accuracy [49].

Understanding sIL-6R response in COVID-19 is of interest given the observations in other inflammatory conditions. Unfortunately, data comparing sIL-6R expression with COVID-19 severity and response are limited. Koutsakos and colleagues found that both IL-6 and sIL-6R concentrations were higher in critically ill patients with COVID-19 compared to patients who were not critically ill [40]. They found that sIL-6R was marginally more accurate than IL-6 at predicting ward versus ICU admission status (AUROC: 0.77 vs 0.7)[40]. In the acute phase of COVID-19, both markers were elevated and exhibited a high degree of within-subject variability compared to healthy patients and those with convalescent disease [40]. Likewise, Moes et al. measured sIL-6R response after tocilizumab administration and noted a rapid increase with a slow decline over 17 days [29]. Based on their population PK model, they estimated that a threshold concentration of 5 μg/mL of tocilizumab would be required to saturate sIL-6R--a value roughly 5-fold higher than in non-COVID-19 patients. However, all patients maintained concentrations above this threshold for over 2 weeks and no difference in tocilizumab PK exposure was discerned between clinical response groups. As a result, uncertainty remains regarding whether sIL-6R monitoring would be useful in evaluating the adequacy of tocilizumab in patients with COVID-19.

Based on available evidence it appears that IL-6 expression in COVID-19 is more similar to that seen in patients with RA versus CRS; that lower concentrations of IL-6 correspond with better clinical outcomes in COVID-19; and that IL-6 expression within the lung is similar between patients with COVID-19 and pneumonia due to other causes. Assuming IL-6 expression is an accurate marker of disease severity in COVID-19, one could argue that disease similarity exists between COVID-19 and other inflammatory conditions. However, IL-6 is an imperfect marker of clinical response in conditions such as RA [40]. Meanwhile, sIL-6R appears to potentially be a more precise measure of host inflammation versus IL-6 [40,48]. Additionally, the pathophysiology driving IL-6 expression in COVID-19 is markedly different (acute viral infection) from conditions like RA, PJIA and SJIA (chronic auto-immune inflammation) and the relationship between tocilizumab treatment and occupancy of its target, IL-6R, is unknown in COVID-19. Therefore, extreme caution is warranted when extrapolating IL-6 and sIL-6R response to COVID-19. Finally, it remains unclear if a causal link exists between IL-6 expression and clinical response in COVID-19.

### Current knowledge of tocilizumab exposure-response for efficacy

There are no PK/PD targets for tocilizumab with respect to its use in chronic or acute inflammatory conditions (e.g., RA, CRS) nor for patients with COVID-19. Without a defined target concentration (e.g., C_max_) or exposure (e.g., AUC), it is challenging to design disease-specific, let alone patient-specific, dosing regimens. To date, it remains unclear whether an exposure threshold exists for tocilizumab in order to confer the benefits observed in randomized trials conducted in patients with COVID-19. As a result, individualized response monitoring (i.e., trending) of clinical response and cautious interpretation of easily obtainable biomarkers (eg., IL-6 and CRP) has become the norm in the clinical setting. The only exposure-response analysis reported to date is provided in the preliminary study by Moes et al. [29]. The investigators predicted individual tocilizumab exposure in 29 patients with severe COVID-19 and computed mean (standard deviation) exposure based on CRP relapse status (post-dose increase in CRP) and survival. They found no differences in exposure between patients with or without CRP relapse nor survivors and non-survivors. These results must be considered exploratory given the limited sample size and single-dose concentration (i.e., all patients received an 8 mg/kg dose).

### Current knowledge of tocilizumab exposure-response for safety

Tocilizumab globally inhibits IL-6 receptors (including both membrane-bound and soluble forms), causing inhibition of potential protective effects of IL-6 and resulting in decreased serum concentrations of acute-phase proteins including CRP [13,35]. While this anti-inflammatory effect can be beneficial in the setting of chronic inflammatory conditions such as RA, global IL-6 inhibition carries the risk of inhibiting an appropriate response to acute infection as well as cell growth and differentiation. Accordingly, adverse events associated with tocilizumab include bacterial infection, intestinal perforation, and pancreatitis [34].

No definitive exposure-toxicity relationship exists for tocilizumab. In the initial FDA review, patients receiving tocilizumab who weighed >100 kg were more likely to experience any infection (41-50%) and serious infections (1.4-4.5%) compared to those who weighed <60 kg (32-38% and 1.3-2.7% respectively). However, the trend towards greater adverse events (including infections) in the >100 kg group was similar between patients who received tocilizumab plus a DMARD compared to those who received DMARD plus placebo [21]. In healthy volunteers, the highest safe and tolerable dose of tocilizumab that did not adversely impact neutrophil counts was 20 mg/kg [27]. The highest C_max_ associated with the maximum tolerated dose (28 mg/kg) in healthy volunteers was 649 μg/mL [27], which is higher than would typically be experienced with a single 8 mg/kg dose. Given the likely increased clearance in critically ill patients with COVID-19, it is unlikely that this exposure would be achieved with a single 8 mg/kg dose. A systematic review and meta-analysis of long-term use in RA patients suggested a possible link with higher doses of tocilizumab (8 mg/kg) to secondary infections, which was less evident in patients receiving lower doses (4 mg/kg) [21,50], but this observation requires further clinical correlation.

### Comparison of fixed and weight-based dosing using Model-informed Simulations

To evaluate the extent to which a fixed dosing approach would yield exposures similar to those experienced by patients enrolled in clinical trials, we conducted focused model-informed dosing simulations. We utilized the distribution of body weight data from patients enrolled in the REMAP-CAP trial to derive a reference range of PK exposures [3,51]. The model structure and parameter estimates were taken from the RA population PK model including both subcutaneous and intravenous administration [25], with linear clearance increased to the value estimated in patient populations with CRS following CAR-T, and used to simulate 100 trials with a sample size equal to the tocilizumab arm of REMAP-CAP (n=353)(51) using NONMEM (version 7.4). Body weights were sampled from a random normal distribution (mean=90.3kg; SD=17.1kg) to reflect the median (IQR) of patients enrolled in the tocilizumab arm of the trial [52].

From the 100 simulated trials, the median (5th, 95th percentiles) of individual exposure measures (i.e., AUC and C_max_) were calculated at the trial level, and the median value of each of these summary statistics across the 100 trials were plotted to define the reference exposure (blue shaded range) in Figure 1. In a separate set of dose simulations, one patient per kg of body weight between 41-160 kg was generated and administered either a fixed or weight-banded dose, as well as a weight-based 8 mg/kg dose. A total of 1000 iterations were performed resulting in 1000 simulated patients at each kg of body weight. Individual simulated exposure measures were calculated for each dosing condition for individual simulated patients, and the median and IQR exposure measures were calculated by binning simulated patients into body weight groups.

Figure 1A and 1B demonstrate that a fixed 400 mg dose results in tocilizumab exposures below those likely achieved in REMAP-CAP where an 8 mg/kg dose was used. A 400 mg fixed-dose is predicted to achieve exposures below the 5th percentile of predicted exposures from REMAP-CAP (8 mg/kg) in excess of 50% of patients weighing more than 66kg. Increasing intra-individual variation in the model (by a factor of four) resulted in a wider reference range (Figure 1C and 1D) and marginally extended the weight range captured by 400 mg fixed-dosing. A higher 600 mg fixed dose (Figure 1E and 1F) results in a greater proportion of simulated patients achieving exposure within the reference range but is still predicted to result in lower than reference exposure in the highest weight stratum.

**Fig. 1a.**
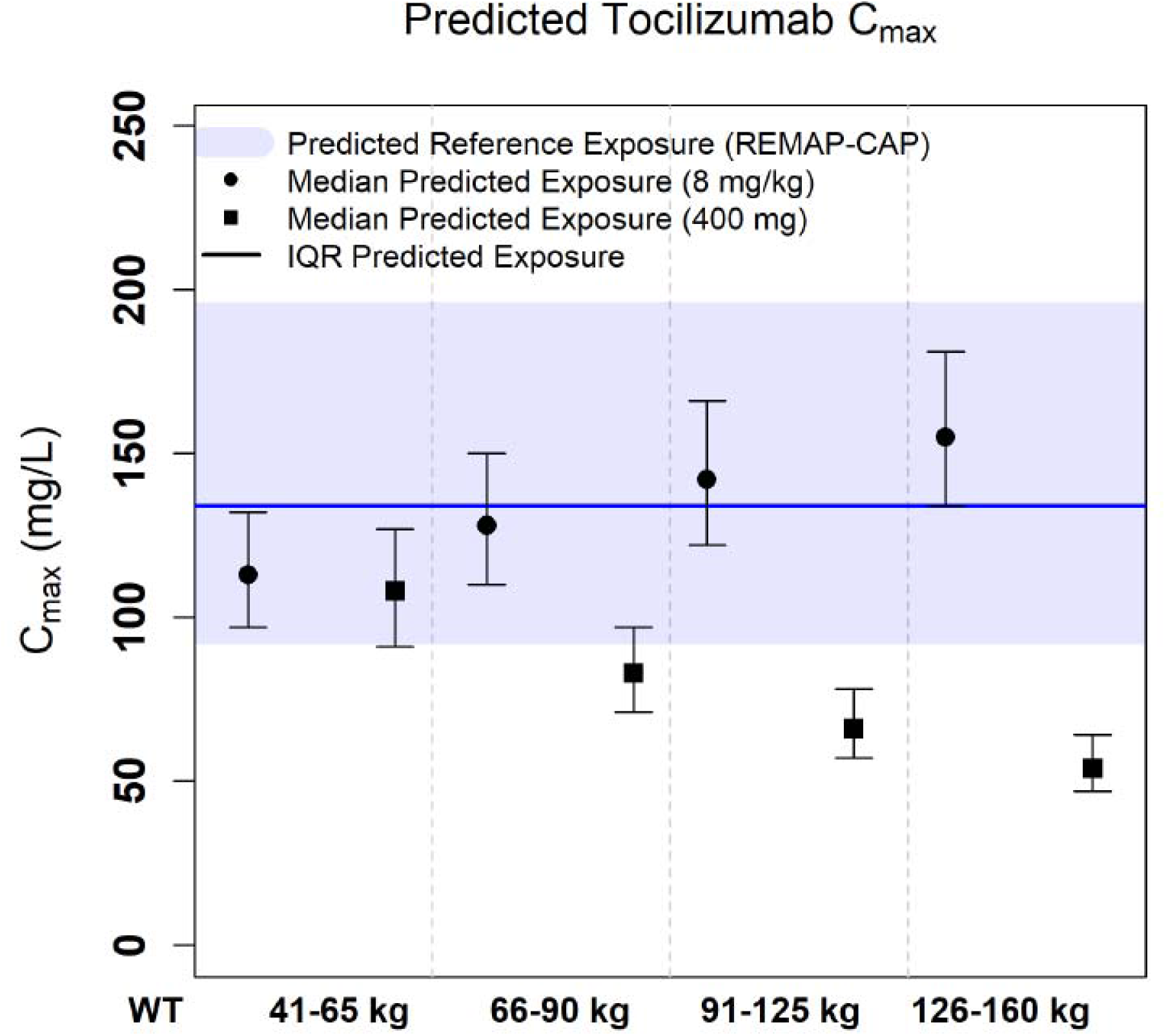
Predicted C_max_ for 400 mg and 8 mg/kg dosing using the reference mode

**Fig. 1b.**
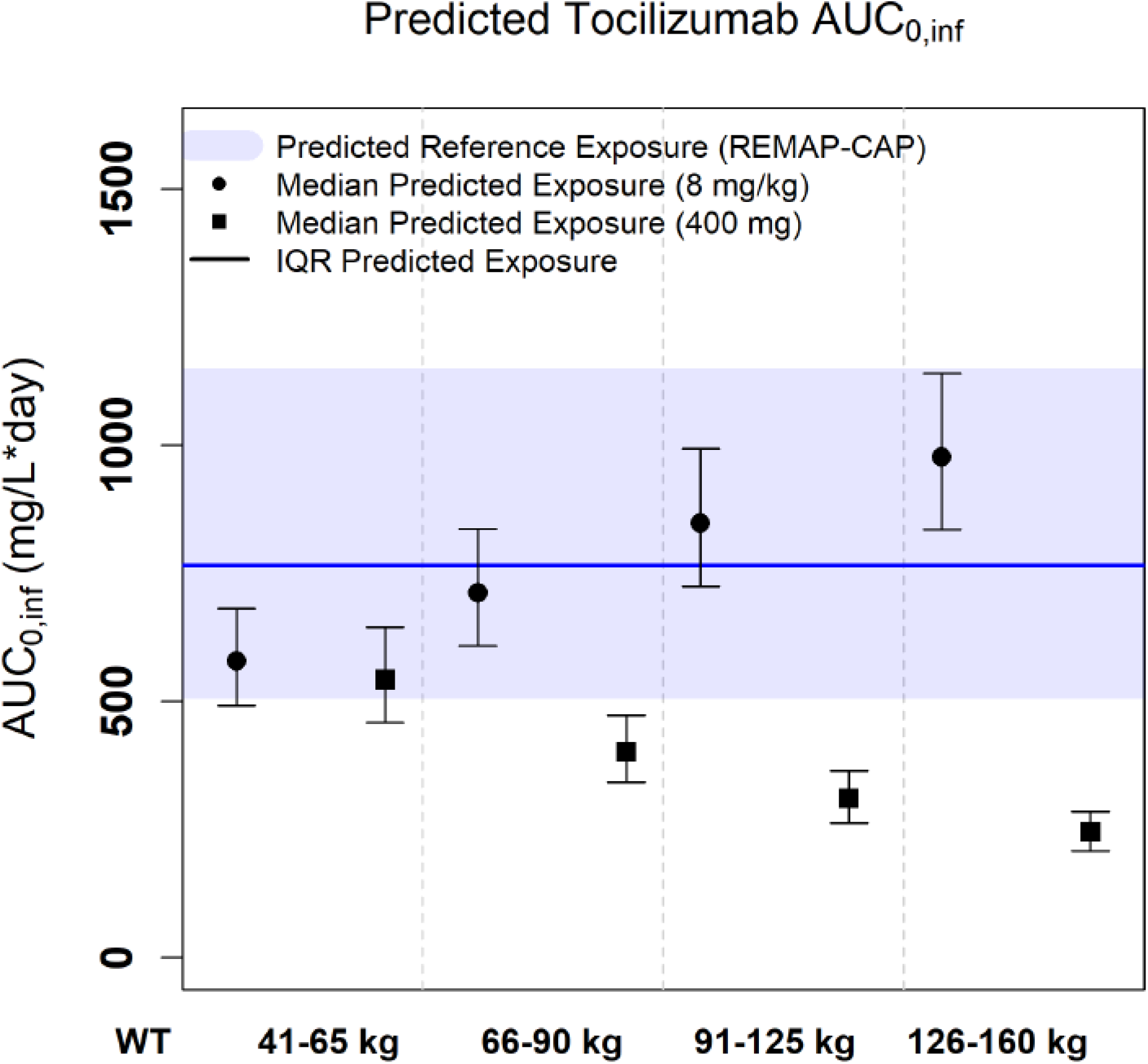
Predicted AUC_(0-inf)_ for 400 mg and 8 mg/kg dosing using the reference model

**Fig. 1c.**
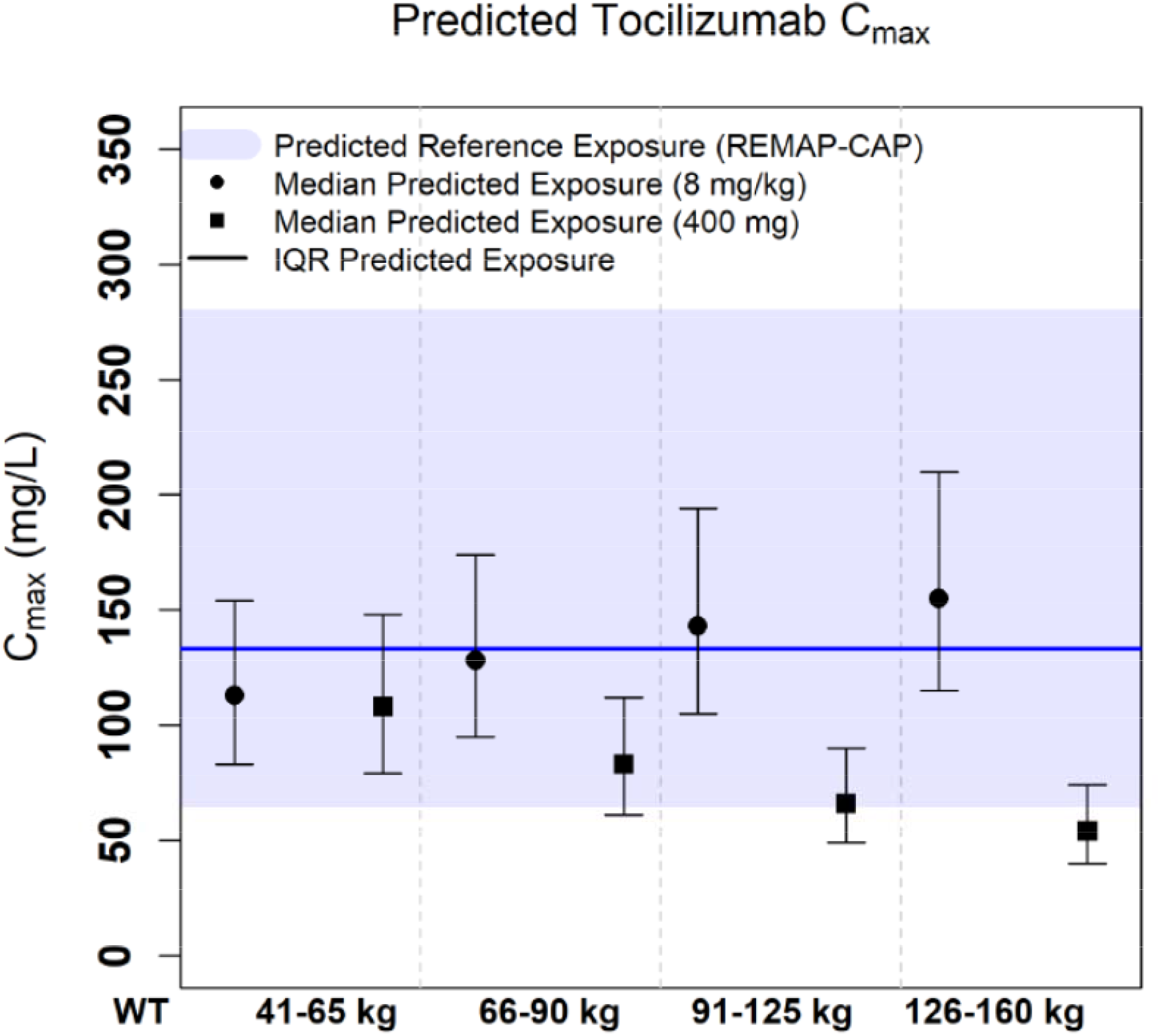
Predicted C_max_ for 400 mg and 8 mg/kg dosing using the reference model with 4-fold inflation of inter-individual variability

**Fig. 1d.**
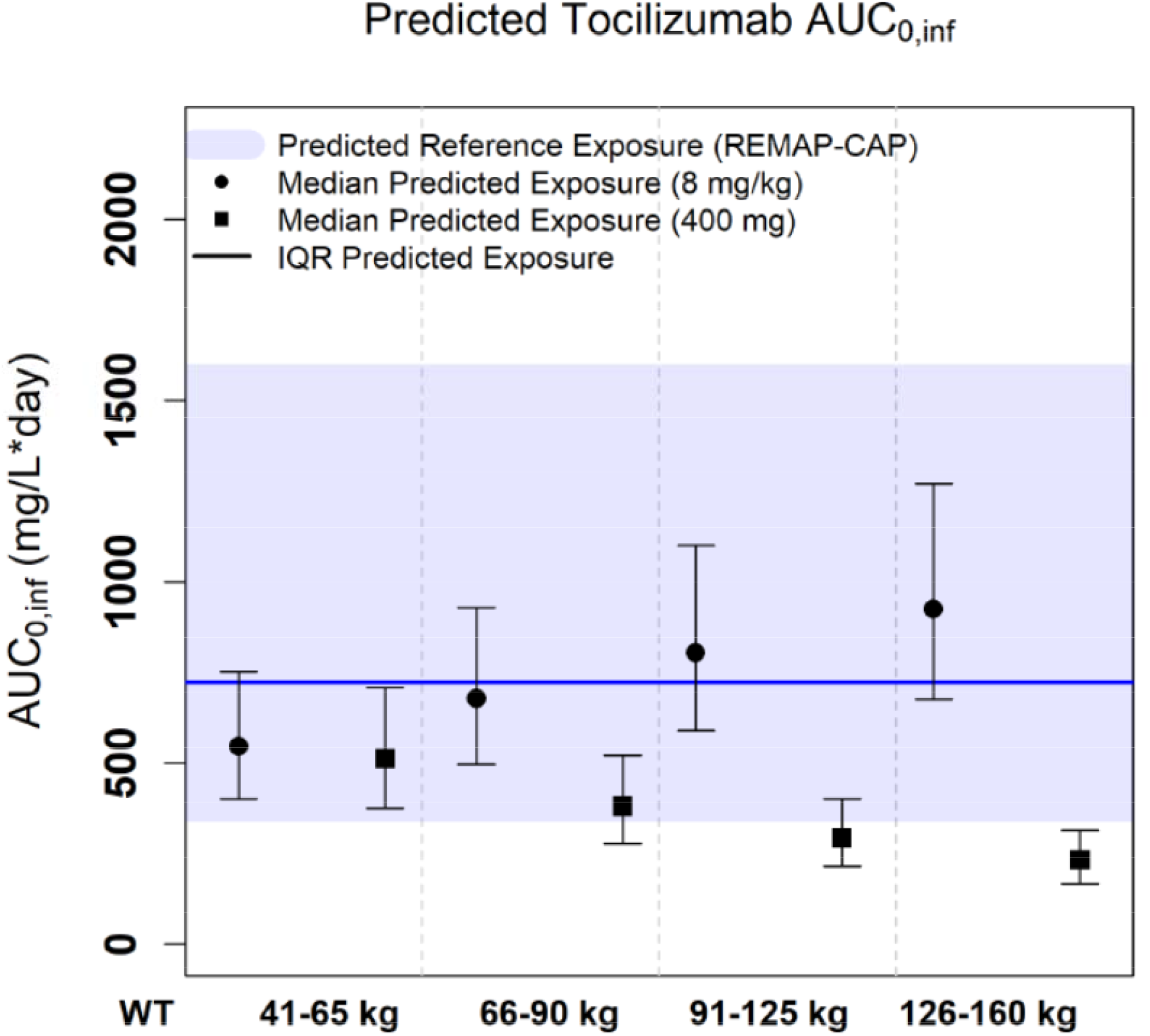
Predicted AUC_(0-inf)_ for 400 mg and 8 mg/kg dosing using the reference model with 4-fold inflation of inter-individual variability

**Fig. 1e.**
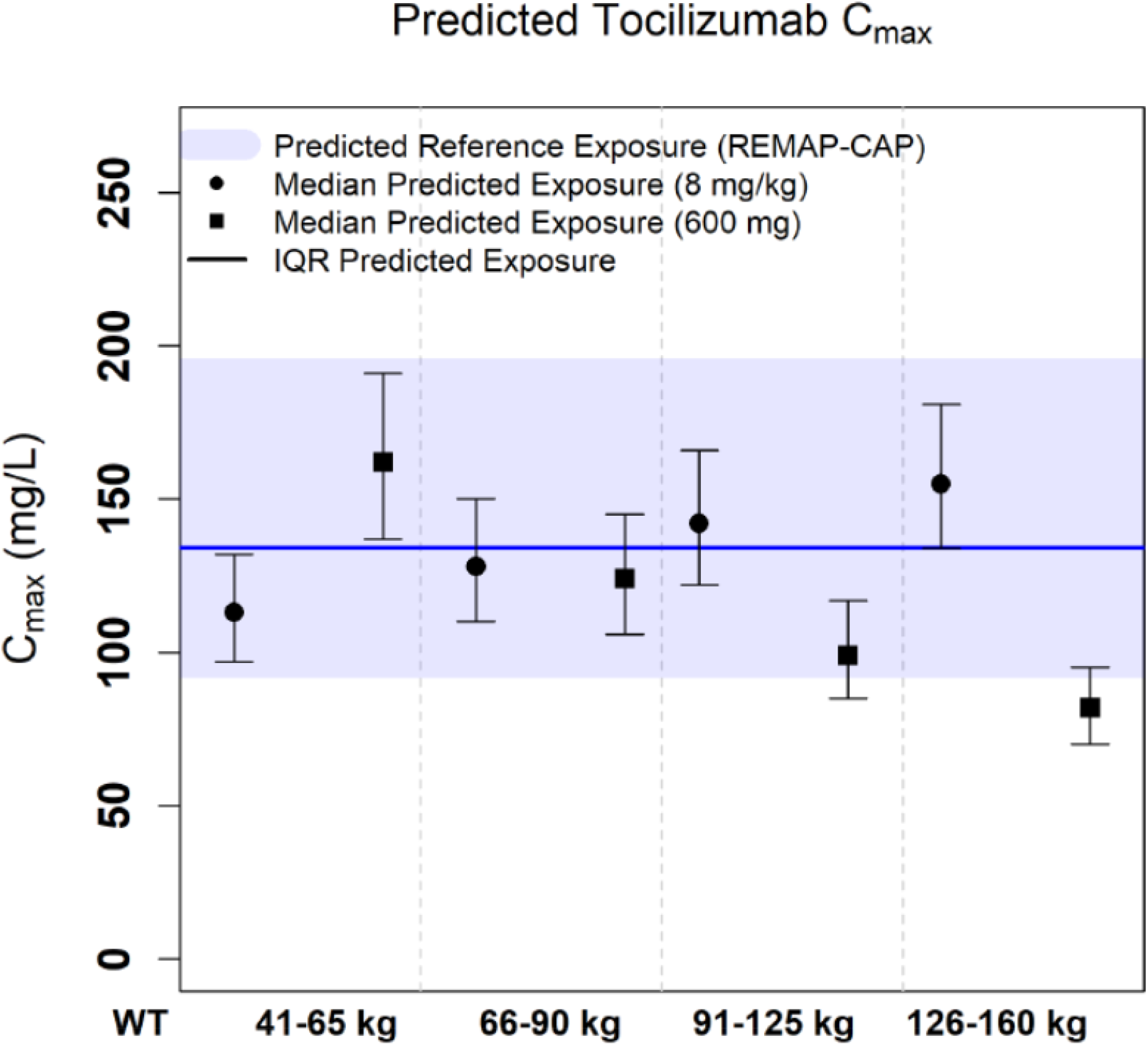
Predicted C_max_ for 600 mg and 8 mg/kg dosing using the reference model

**Fig. 1f.**
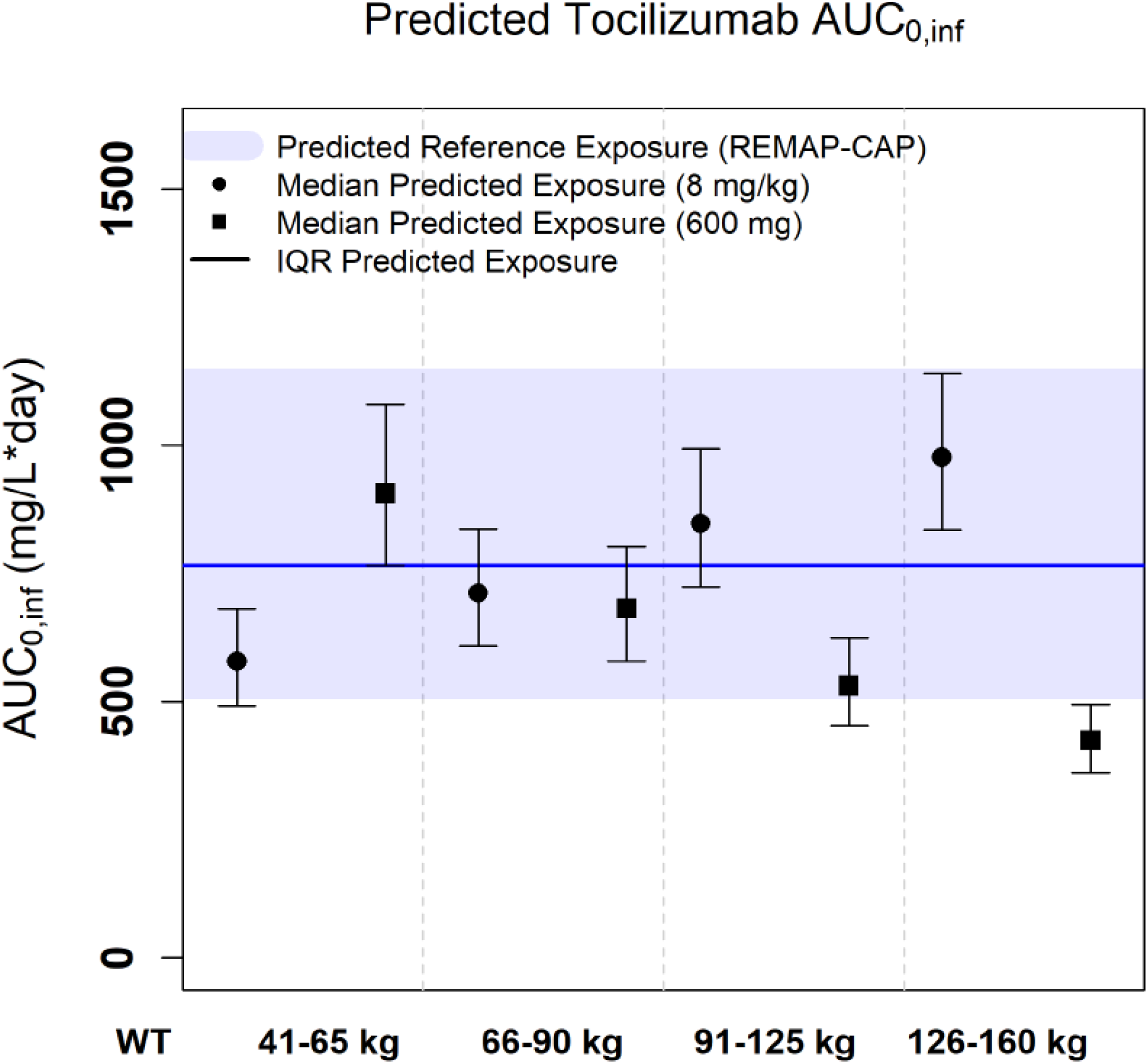
Predicted AUC_(0-inf)_ for 600 mg and 8 mg/kg dosing using the reference model

Notably, across all of these simulation conditions the 8 mg/kg dosing arm is predicted to result in increasing exposure with increasing body weight, such that exposure is not matched across body weight groups. A weight-banded dosing approach with less frequent dose adjustments than RECOVERY and using practical 200 mg dosing increments is predicted to result in improved exposure-matching across the body weight range while still achieving exposures within the reference range predicted for REMAP-CAP (Figure 1G and 1H).

**Fig. 1g.**
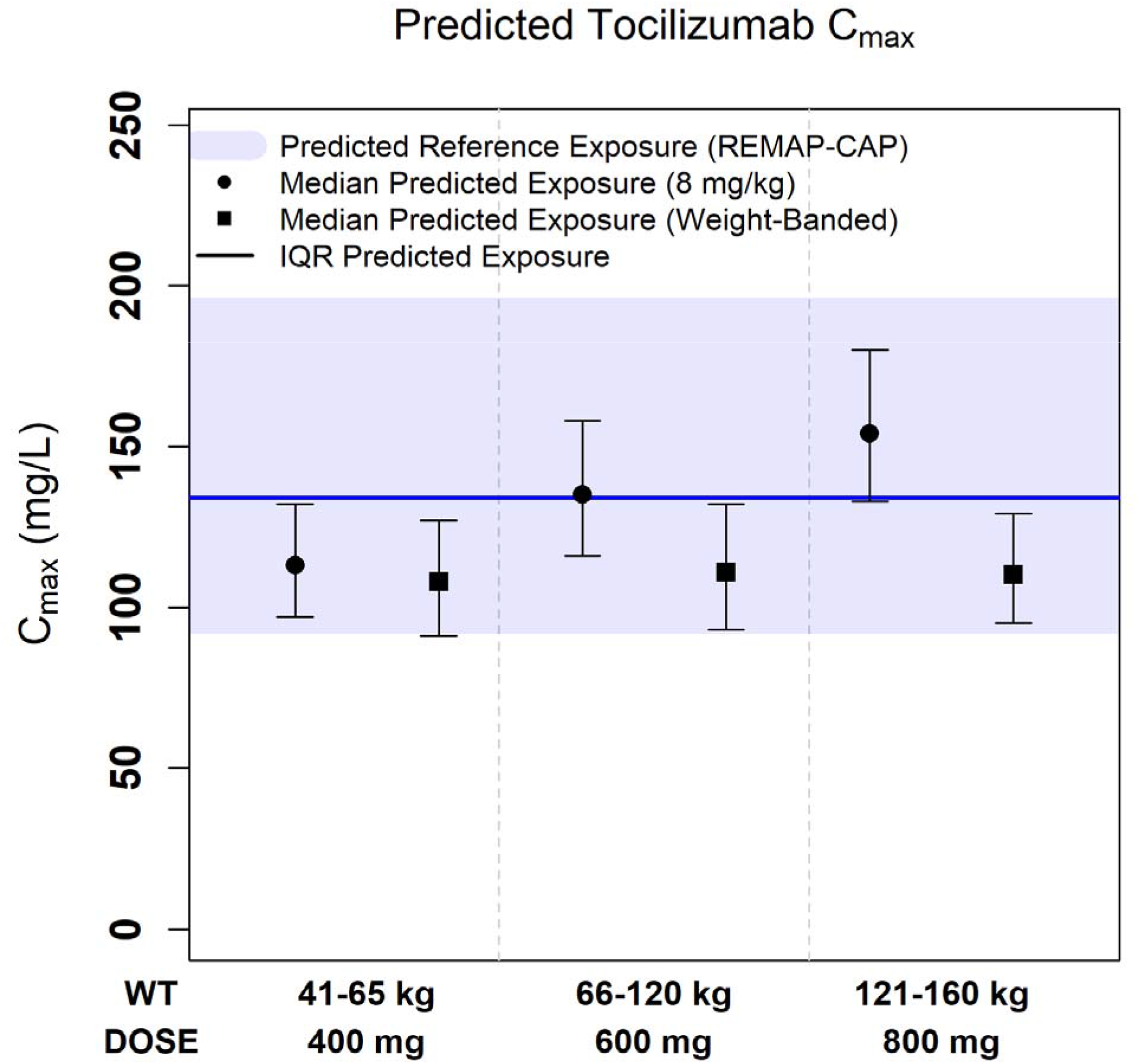
Predicted C_max_ for weight-banded fixed dosing and 8 mg/kg dosing using the reference model

**Fig. 1h.**
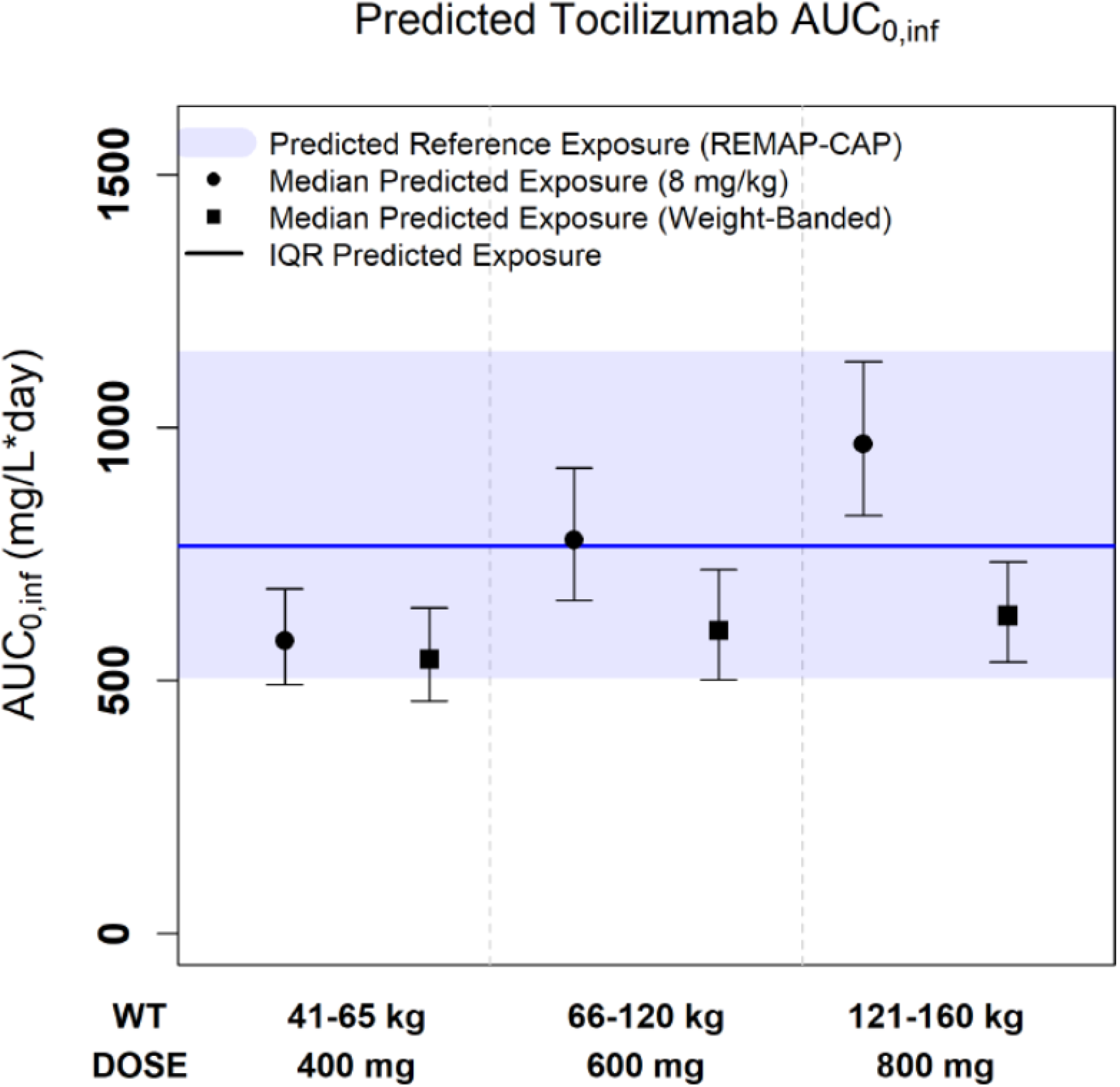
Predicted AUC_(0-inf)_ for weight-banded fixed dosing and 8 mg/kg dosing using the reference model

## Discussion

The optimal tocilizumab exposure in patients with COVID-19 is not known. We sought to identify a fixed dosing regimen that is predicted to achieve exposures similar to the weight-based dosing regimens used in REMAP-CAP with the aim of informing drug conservation strategies [3]. We were not able to predict exposures from RECOVERY, because patient body weight distributions were not provided in that trial [4]. Our simulations suggest that patients weighing ≥90 kg are unlikely to achieve exposures that fall within the reference range based upon the predicted exposures within REMAP-CAP, even if a 600 mg fixed-dose was applied. Additionally, our analysis found that a weight-banded approach, similar to the dosing strategy employed in RECOVERY but with more conservative dose adjustments (using practical 200 mg increments and alternate weight bands), yielded exposures that fell within the 90% predictive interval of exposures from REMAP-CAP. Our findings support exploration of an alternate weight-banded dosing strategy, but such dosing will require clinical validation.

On the other hand, our simulation results may not fully generalize to critically ill COVID-19 patients. It remains unclear whether the tocilizumab exposures approximated from the large COVID-19 platform trials are necessary, given the relatively low drug concentrations required to saturate the sIL-6R signaling pathway --a pathway which appears potentially more predictive of clinical outcomes than IL-6 alone. It is known that sIL-6R levels are significantly higher in critically ill COVID-19 patients compared to non-critically ill COVID-19 patients and healthy volunteers, which may result in more rapid target-mediated tocilizumab clearance [40]. Furthermore protein and IgG catabolism has been shown to be increased in critically ill patients in general. This may partly explain the lower exposures seen in patients with CAR T therapy-associated CRS compared to patients with PJIA [43]. Of note, albumin and total protein were also modifiers of tocilizumab clearance in the final tocilizumab PK model [21]. Additionally, corticosteroids, which are now the standard of care in all COVID-19 patients who require supplemental oxygen, are associated with upregulation of IL-6R (including sIL6-R)[53,54]. Such increased expression could also contribute to enhanced target-mediated clearance and could reduce concentrations below saturation levels sooner. Interestingly, although IL-6 is generally not elevated during pregnancy, sIL-6R concentrations are higher in pregnant women compared to non-pregnant women [55]. The increase in blood volume associated with pregnancy may further reduce tocilizumab concentrations necessitating larger doses; additional studies are needed in pregnant patients. Although fixed doses appear to produce more homogenous PK exposures in a small number of patients with COVID-19 [29], there is likely to be higher between-patient variability in drug exposure among critically ill patients. Thus, more work is needed to understand the clinical PK of tocilizumab in COVID-19.

We look forward to further information to inform tocilizumab PK/PD in COVID-19 disease from the COVACTA study, which used weight based dosing of tocilizumab 8 mg/kg and planned to collect of serum concentrations of IL-6, sIL-6R, and CRP [56]. Limited observational data suggest that the therapeutic effect of tocilizumab in COVID-19 is not compromised when using lower (40-200 mg)[57], or fixed dose of tocilizumab (400 mg)[11,58]. However, randomized controlled trials have not yet evaluated this approach.

Higher doses of IV tocilizumab chosen for RA allow for extended interval dosing in the outpatient setting (i.e., every 4 weeks), which may not be necessary in an acute condition such as COVID-19. Given that sIL-6R saturation appears to be achieved with relatively low tocilizumab concentrations (1 to 5 μg/mL) [29,35], it is possible that lower doses could be as effective as higher doses for the treatment of COVID-19 [13,21]. Additionally, it has been suggested that the relative lower IL-6 levels in COVID-19 as compared to CRS may potentially support consideration of lower IL-6R antagonist doses in response to lower cytokine concentrations [13]. A lower tocilizumab dose that achieves receptor saturation but for a shorter duration could also theoretically minimize the risk of secondary opportunistic infections associated with prolonged IL-6 blockade without compromising efficacy.

Finally, due to the lack of objective targets, the adaptive trials conducted to date have allowed re-dosing of tocilizumab at the discretion of treating clinicians, largely based on clinical response [3,4]. Given the expected half-life of tocilizumab ranging from 11 to 13 days, additional doses of this therapy based on clinical response are likely unnecessary in COVID-19; avoiding second doses could further reserve its limited supply [9,10,23].

## Conclusion

In the treatment of COVID-19, it is currently unclear whether weight-based dosing provides advantages over a fixed dosing strategy, as the largest published trials to date establishing the benefit of tocilizumab in COVID-19 have used weight-based dosing and/or dose banding strategies. Fixed tocilizumab dosing of 400 mg across body weight strata does not appear to approximate the tocilizumab exposures (i.e., AUC) estimated from clinical trials showing benefit. Currently, a dose-exposure-response relationship for tocilizumab in the treatment of COVID-19 is not yet described, and there remains uncertainty regarding the extent to which COVID-19 may alter individual PK parameters. The use of a fixed dosing regimen may reduce the probability of “underdosing” in some patients whereas the risk of “overexposure” with weight-based dosing in high weight patients is non-trivial. To date, the clinical contribution of “overexposure” on the risk of secondary infections is unclear. Since an established PK/PD relationship for tocilizumab is not known, it is reasonable for drug dosing to target an exposure matching approach across individuals in the target population [14]. It is reasonable to consider a fixed dose approach in the setting of critical limitations in drug supply, in an attempt to treat as many patients as possible who would qualify for tocilizumab therapy.

We recommend that if jurisdictions or individual hospital sites move to adopt alternate dosing approaches (a likely possibility in the scenario where ethical rationing of limited drug product is in effect), it is studied - ideally with real-time monitoring - to provide data on patient centered outcomes and the real-world effect of fixed dose, or lower-dose tocilizumab therapy in COVID-19 disease. In addition, the current landscape of clinical trials is rapidly evolving, and adaptive trials have proven especially useful during this pandemic due to their agile and flexible design in exploring repurposed therapies. We highlight here the importance of rational dosing of medications through PK/PD, and strongly advocate that current and future trials incorporate PK/PD considerations into their trial designs in order to better inform optimized dosing strategies at the bedside.

## Data Availability

Data sharing not applicable to this article as no datasets were generated or analyzed during the current study (this is a review article, not a primary study)

## Declarations

### Funding

No institutional or corporate funding was received by any authors for this work.

### Conflicts of Interest/Competing Interests

All authors do not declare any conflict of interest including pertinent commercial and other relationships related to this work. SCJJ has previously received a presenter’s honorarium from Sunovion, unrelated to the present work. EL has previously received a presenter’s honorarium from the Ontario Pharmacists’ Association, unrelated to the present work. NJR reports honoraria from ASHP and receipt of research funds from AACP and Paratek, unrelated to the present work. SR has previously received a presenter’s honorarium from Merck, unrelated to the present work.

### Code Availability

Not applicable (this is a review article, not a primary study) Modeling software: NONMEM (Version 7.4)

### Plotting software

R (Version 3.6.3)

### Authors’ Contributions

EL wrote the first draft of the manuscript. EL, SR, and BL contributed to the conception of the manuscript. NJR and RC provided methodology support and performed PK/PD modeling. All authors (EL, SJ, SR, BL, RLC, WJM, NJR) revised it critically for important intellectual content, and approved the final version.

### Ethics Approval/Consent to Participate/Consent for Publication

Not applicable (review article)

## Acknowledgements

We would like to acknowledge Dr. Srinisvas Murthy (University of British Columbia) and the REMAP-CAP trial team for providing us with the weight distributions of the study population in the REMAP-CAP study for our pharmacokinetic modeling analysis.

## Notes

### Author Declarations

Not applicable (review article/simulation).

